# Levels of SARS-CoV-2 population exposure are considerably higher than suggested by seroprevalence surveys

**DOI:** 10.1101/2021.01.08.21249432

**Authors:** Siyu Chen, Jennifer A Flegg, Lisa J White, Ricardo Aguas

## Abstract

Accurate knowledge of accurate levels of prior population exposure has critical ramifications for preparedness plans of subsequent SARS-CoV-2 epidemic waves and vaccine prioritization strategies. Serological studies can be used to estimate levels of past exposure and thus position populations in their epidemic timeline. To circumvent biases introduced by decaying antibody titers over time, population exposure estimation methods should account for seroreversion, to reflect that changes in seroprevalence measures over time are the net effect of increases due to recent transmission and decreases due to antibody waning. Here, we present a new method that combines multiple datasets (serology, mortality, and virus positivity ratios) to estimate seroreversion time and infection fatality ratios and simultaneously infer population exposure levels. The results indicate that the average time to seroreversion is six months, and that true exposure may be more than double the current seroprevalence levels reported for several regions of England.

## Introduction

The COVID-19 pandemic has inflicted devastating effects on global populations and economies^1^. Levels and styles of reporting epidemic progress vary considerably across countries^2^, with cases consistently being under-reported and case definitions changing significantly over time. Therefore, the scientific and public health communities turned to serological surveys as a means to position populations along their expected epidemic timeline, and thus provide valuable insights into COVID-19 lethality ^3,4^. Those prospects were frustrated by apparent rapid declines in antibody levels following infection^5-7^. Population wide antibody prevalence measurements can significantly underestimate the amount of underlying population immunity with obvious implications for intervention strategy design and vaccine impact measurement.

Continued research efforts to determine the correlates for protective immunity against disease and infection have found that while antibody titers are poor indicators of sustained immunity, cellular immunity can play a determinant role in limiting susceptibility to further SARS-CoV-2 challenges in previously exposed individuals^8,9^. Unfortunately, performing T cell assays at scale is technically challenging and expensive, which justified the decision to conduct a series of serology surveys (some of which are still underway) in many locations globally to provide a better understanding of the extent of viral spread among populations^10^.

In England, a nationwide survey sampling more than 100,000 adults was performed from 20 June to 13 July 2020. The results suggested that 13% and 6% of the population of London and England, respectively, had been exposed to SARS-CoV-2, giving an estimated overall infection fatality ratio (IFR) of 0.90%^11^. Although corrections were made for the sensitivity and specificity of the test used to infer seroprevalence, declining antibody levels were not accounted for. This is a limitation of the approach, potentially resulting in underestimates of the true levels of population exposure^12^ and an overestimate of the IFR.

We now have a much clearer picture of the time dynamics of humoral responses following SARS-CoV-2 exposure, with antibody titers remaining detectable for approximately 6 months^13,14^. Commonly used serological assays have a limit of antibody titer detection, below which a negative result is yielded. Hence, a negative result does not necessarily imply absence of antibodies, but rather that there is a dynamic process by which production of antigen targeted antibodies diminishes once infection has been resolved, resulting in decaying antibody titers over time. As antibody levels decrease beyond the limit of detection, seroreversion occurs.

We define the seroreversion rate as the inverse of the average time taken following seroconversion for antibody levels to decline below the cut-off for testing seropositive. In a longitudinal follow-up study, antibodies remained detectable for at least 100 days^6^. In another study^15^, seroprevalence declined by 26% in approximately three months, which translates to an average time to seroreversion of around 200 days. However, this was not a cohort study, so newly admitted individuals could have seroconverted while others transitioned from positive to negative between rounds, leading to an overestimation of the time to seroreversion.

Intuitively, if serology were a true measure of past exposure, we would expect a continually increasing prevalence of seropositive individuals over time. However, data suggest this is not the case^16^, with most regions in England showing a peak in seroprevalence at the end of May 2020. This suggests seroreversion plays a significant role in shaping the seroprevalence curve and that the time since the first epidemic peak will influence the extent to which subsequent seroprevalence measurements underestimate the underlying population attack size (proportion of the population exposed). We argue that the number of people infected during the course of the epidemic can be informed by data triangulation, i.e., by combining numbers of deceased and seropositive individuals over time. For this linkage to be meaningful, we need to carefully consider the typical SARS-CoV-2 infection and recovery timeline (Figure 1).

Most individuals, once infected, experience an incubation period of approximately 4.8 days (95% confidence interval (CI): 4.5–5.8)^17^, followed by the development of symptoms, including fever, dry cough, and fatigue, although some individuals will remain asymptomatic throughout. Symptomatic individuals may receive a diagnostic PCR test at any time after symptoms onset; the time lag between symptoms onset and date of test varies by country and area, depending on local policies and testing capacity. Some individuals might, as their illness progresses, require hospitalization, oxygen therapy, or even intensive care, eventually either dying or recovering.

The day of symptoms onset, as the first manifestation of infection, is a critical point for identifying when specific events occur relative to each other along the infection timeline. The mean time from symptoms onset to death is estimated to be 17.8 days (95% credible interval (CrI): 16.9–19.2) and to hospital discharge 24.7 days (22.9–28.1)^18^. The median seroconversion time for IgG (long-lasting antibodies thought to be indicators of prior exposure) is estimated to be 14 days post-symptom onset; the presence of antibodies is detectable in less than 40% of patients within 1 week of symptoms onset, rapidly increasing to 79.8% (IgG) at day 15 post-onset^19^. We assume onset of symptoms occurs at day 5 post-infection and that it takes an average of 2 additional days for people to have a PCR test. Thus, we fix the time lag between exposure and seroconversion, *δ*_*ϵ*_, at 21 days, the time lag between a PCR test and death, *δ*_*P*_, as 14 days, and assume that seroconversion in individuals who survive occurs at approximately the same time as death for those who don’t (Figure 1).

**Figure 1.**
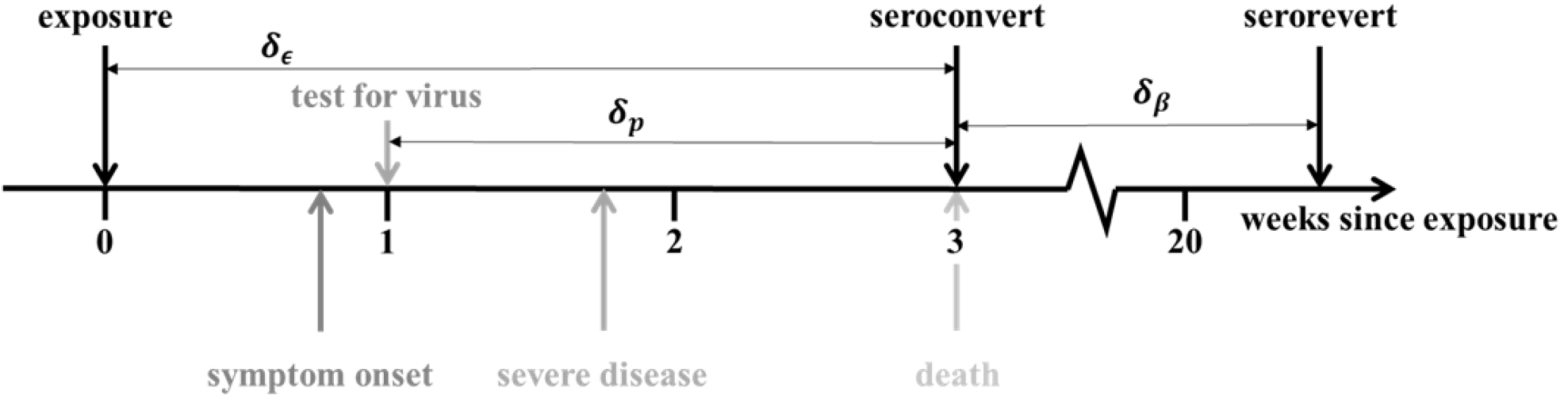
Progression of exposed individuals through the various clinical (below the timeline), and diagnostic (above the timeline) stages of infection and recovery. Stages marked in grey represent events that may happen, with a probability consistent with the darkness of the shade of grey.

Thus, we propose to use population level dynamics (changes in mortality and seroprevalence over time) to estimate three key quantities: the seroreversion rate, the IFR, and the total population exposure over time. We developed a Bayesian inference method to estimate said quantities, based on official epidemiological reports and a time series of serology data from blood donors in England, stratified by region^16^ – see Materials and Methods for more details. This dataset informed the national COVID-19 serological surveillance and its data collection was synchronous with the “REACT” study^11^. The two sero-surveys use different, but comparable, antibody diagnostic tests^20^. While “REACT” used a lateral flow immunoassay (LFIA) test for IgG^11^, the data presented here were generated using the Euroimmun® assay. The independent “REACT” study acts as a validation dataset, lending credence to the seroprevalence values used. For example, seroprevalence in London was reported by REACT to be 13.0% (12.3–13.6) for the period 20 June to 13 July 2020. In comparison, the London blood-donor time series indicated seroprevalence to be 13.3% (8.4–16) on 21 June 2020.

We developed a method that combines daily mortality data with seroprevalence in England, using a mechanistic mathematical model to infer the temporal trends of exposure and seroprevalence during the COVID-19 epidemic. We fit the mathematical model jointly to serological survey data from seven regions in England (London, North West, North East (North East and Yorkshire and the Humber regions), South East, South West, Midlands (East and West Midlands combined), and East of England) using a statistical observation model. For more details on the input data sources, mechanistic model and fitting procedure, see the Materials and Methods section. We considered that mortality is perfectly reported and proceeded to use this anchoring variable to extrapolate the number of people infected 3-weeks prior. We achieved this by estimating region-specific IFRs (defined as *γ*_*i*_), which we initially assumed to be time invariant, later relaxing this assumption. The identifiability of the IFR metric was guaranteed by using the serological data described above as a second source of information on exposure. From the moment of exposure, individuals seroconvert a fixed 21 days later and can then serorevert at a rate, β, that is estimated as a global parameter. We thus have both mortality and prevalence of seropositivity informing SARS-CoV-2 exposure over time.

Several other research groups have used mortality data to extrapolate exposure and as a result provide estimates for IFR. Some IFR estimates were published assuming serology cross-sectional prevalence to be a true reflection of population exposure, while others used infection numbers generated by mechanistic dynamic models fit to mortality data^21^. Most recently, sophisticated statistical techniques have been used, which take into account the time lag between exposure and seroconversion when estimating the underlying population exposure from seroprevalence measurements^22^, with one study also considering seroreversion^23^. Our method is very much aligned with the latter but is applied at a subnational level while using a dataset that has been validated by an independent, largely synchronous study.

## Results

Results from the fixed IFR inference method show excellent agreement with serological data (Figure 2). We found that, after seroconverting, infected individuals remain seropositive for about 176 days on average (95% CrI: 159-197) (Table 1, Table S1, and Figure 2—figure supplement 1).

**Table 1.**
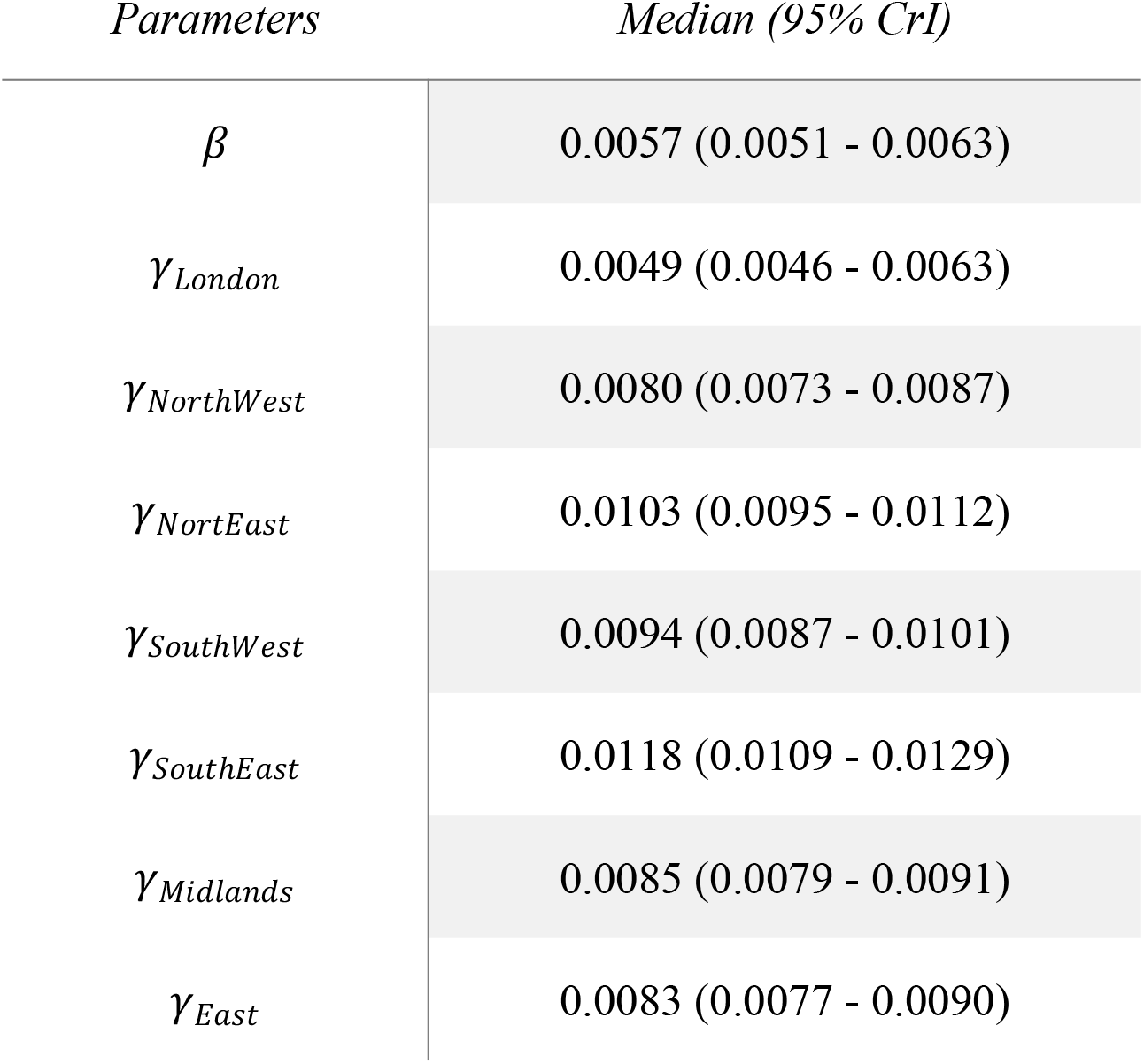
Marginal median parameter estimates and 95% CrI for the constant IFR model. β is the rate of seroreversion and *γ* denotes the IFR. The estimated median time to seroreversion given by 1 /*β* is 176 (95% CrI: 159-197 days)

| *Parameters* | *Median (95% CrI)* |
| --- | --- |
| $\beta$ | 0.0057 (0.0051 - 0.0063) |
| $\gamma_{London}$ | 0.0049 (0.0046 - 0.0063) |
| $\gamma_{NorthWest}$ | 0.0080 (0.0073 - 0.0087) |
| $\gamma_{NorthEast}$ | 0.0103 (0.0095 - 0.0112) |
| $\gamma_{SouthWest}$ | 0.0094 (0.0087 - 0.0101) |
| $\gamma_{SouthEast}$ | 0.0118 (0.0109 - 0.0129) |
| $\gamma_{Midlands}$ | 0.0085 (0.0079 - 0.0091) |
| $\gamma_{East}$ | 0.0083 (0.0077 - 0.0090) |

**Figure 2.**
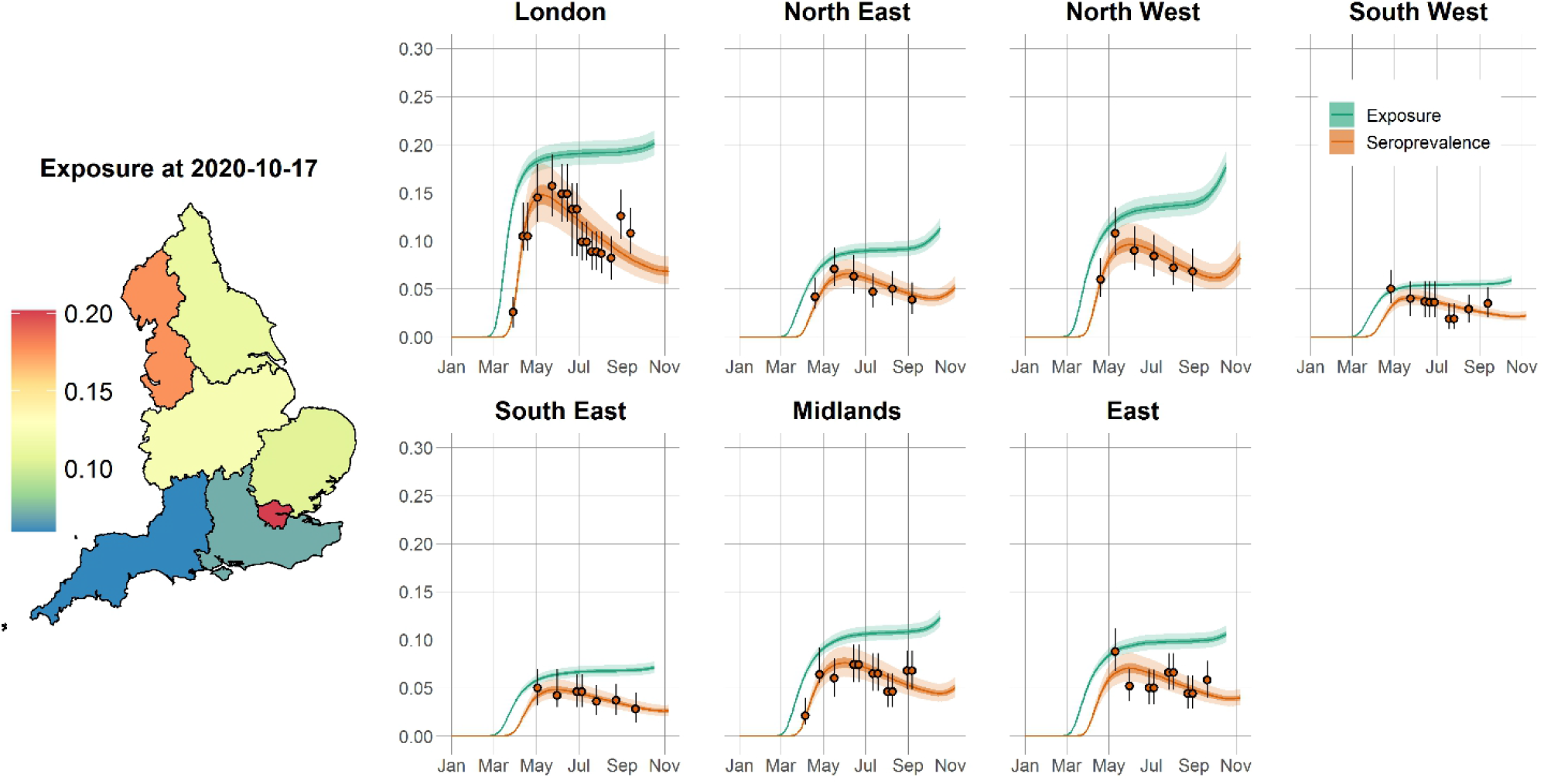
Time course of the SARS-CoV-2 pandemic up to 7 November 2020 for seven regions in England. The solid orange circles and black error bars in each regional panel represent the observed seroprevalence data and their confidence interval, respectively, after adjusting for the sensitivity and specificity of the antibody test. The green and orange lines show the model predictions of median exposure and seroprevalence, respectively, while the shaded areas correspond to 95% CrI. The regional predicted exposure levels (expressed as the proportion of the population that has been infected) as of 17 October 2020 are shown on the map of England.

This relatively rapid (approximately six months) seroreversion is similar to other estimates from experimental studies^13,14^, and might explain the reported 83% protection against reinfection within 6 months of disease in UK patients^24^. As a consequence of this rapid seroreversion, epidemic progression will result in an increasing gap between measured serology prevalence levels and cumulative population exposure to the virus. Ultimately, this may mean that more than twice as many people have been exposed to the virus relative to the number of people who are seropositive (Figure 2), raising questions about the relevance of serological data for informing policy decisions moving forward. We also estimated age-independent IFRs for seven English regions (means ranging from 0.49% to 1.18% - Table 1) that are in very good agreement with other estimates for England^25^.

The estimated IFRs are noticeably lower for London, which can be explained by differences in population age structure across the seven regions considered here. London has a considerably younger population than other regions of England, which, associated with increasing severity of disease with increasing age^26,27^, results in a lower expected number of fatalities given a similar number of infections. This could be construed as a possible explanation for fluctuations in estimates of the country-wide IFR over time^25^, as outbreaks occur intermittently across regions with different underlying IFRs. An alternative interpretation of IFR trends in England is that individuals who are more likely to die from infection (due to some underlying illness or other risk factors) will do so earlier. This means that as the epidemic progresses, a selection (through infection) for a decrease in average population frailty (a measure of death likelihood once infected) is taking place and, consequently, a reduction in the ratio of deaths to infections. To test this hypothesis, we constructed an alternative formulation of our modelling approach, whereby the IFR at a specific time is dependent on the stage of epidemic progression – Figure 3. It is extremely difficult to extrapolate the underlying risk of infection from reported case data due to the volatility in testing capacity. Hence, we propose that the optimal metric for epidemic progression is the cumulative test positivity ratio. In the absence of severe sampling biases, the test positivity ratio is a good indicator of changes in underlying population infection risk, as a larger proportion of people will test positive if infection prevalence increases. In fact, it is clear from (Figure 3 – figure supplement 3 that the test positivity ratio is a much better indicator of exposure than the case fatality ratio (CFR) or the hospitalization fatality ratio (HFR), since it mirrors the shape of the mortality incidence curve. For the time-varying IFR, we took the normalized cumulative test positivity ratio time series and applied it as a scalar of the maximum IFR value estimated for each region – for more details can be found in the Materials and Methods section.

**Figure 3.**
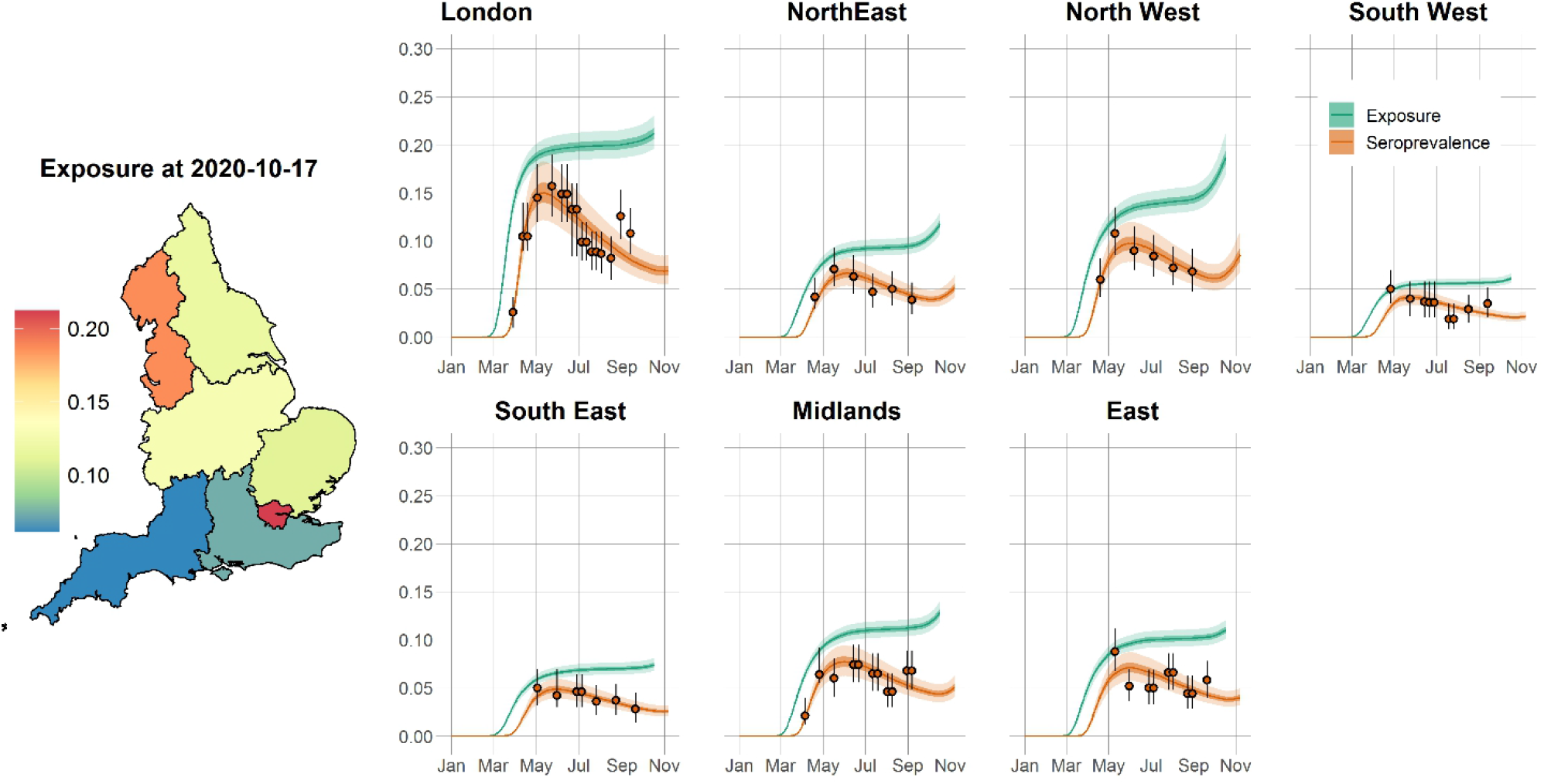
Time course of the SARS-Cov-2 pandemic up to 7 November 2020 for seven regions in England for time-varying IFR model. The orange solid circles and black error bars in each regional panel represent the observed seroprevalence data and its confidence interval after adjusting for the sensitivity and specificity of the antibody test. The green and orange lines show the median time-varying IFR model predictions for exposure, and seroprevalence, respectively, while the shaded areas correspond to 95% CrI. The regional median predicted exposure levels (expressed as the proportion of the population that has been infected) as of 17 October 2020 are shown on the map of England.

Results from the time-varying IFR model indicate that the population of London might have undergone a significant frailty selection process during the first wave of the epidemic and now shows a significantly lower IFR compared with March/April 2020 (Figure 3 – figure supplement 2). Interestingly, no statistically significant time-dependence on IFR was inferred for any of the remaining regions (Figure 3 – figure supplement 1), suggesting this phenomenon is dependent on age structure.

We can eliminate exposure levels as the main driver of this process as there is no clear temporal signal for IFR for the only other region (North West) with a comparable force of selection (i.e., similar predicted exposure levels). It seems that in younger populations, with a lower subset of very frail individuals, this selection will be more pronounced. Overall, the estimates obtained with both models are very consistent, with the estimated credible intervals for the time varying IFR model including the median estimated obtained for the fixed IFR model (Figure 3 – figure supplement 1 and Table S3).

## Discussion

Given the current polarization of opinion around COVID-19 natural immunity, we realize that our results are likely to be interpreted in one of two conflicting ways: (1) the rate of seroreversion is high, therefore achieving population (herd) immunity is unrealistic, or (2) exposure in more affected places such as London is much higher than previously thought, and population immunity has almost been reached, which explains the decrease in IFR over time. We would like to dispel both interpretations and stress that our results do not directly support either. Regarding (1), it is important to note that the rate of decline in neutralizing antibodies, reflective of the effective immunity of the individual, is not the same as the rate of decline in seroprevalence. Antibodies may visibly decline in individuals yet remain above the detection threshold for antibody testing^6^. Conversely, if the threshold antibody titer above which a person is considered immune is greater than the diagnostic test detection limit, individuals might test positive when in fact they are not effectively immune. The relationship between the presence and magnitude of antibodies (and therefore seropositive status) and protective immunity is still unclear, with antibodies that provide functional immunity only now being discovered^13^. Furthermore, T cell mediated immunity is detectable in seronegative individuals and is associated with protection against disease^8^. Therefore, the immunity profile for COVID-19 goes beyond the presence of a detectable humoral response. We believe our methodology to estimate total exposure levels in England offers valuable insights and a solid evaluation metric to inform future health policies (including vaccination) that aim to disrupt transmission. With respect to (2), we must clarify that decreasing IFR trends can result solely from selection processes operating at the intersection of individual frailty and population age structure. Likewise, the lower IFR in London can be attributed to its relatively younger population when compared with populations in the other regions of England. In conclusion, a method that accounts for seroreversion using mortality data allows the total exposure to SARS-CoV-2 to be estimated from seroprevalence data. The associated estimate of time to seroreversion of 176 days (95% CrI: 159-197) lies within realistic limits derived from independent sources. The total exposure in regions of England estimated using this method is more than double the last seroprevalence measurements. Implications for the impact of vaccination and other future interventions depend on the, as yet uncharacterized, relationships between exposure to the virus, seroprevalence, and population immunity. To assess vaccination population impact one can consider the population at risk to be those individuals who are seronegative, those with no past exposure (confirmed or predicted), or those with no T cell reactivity. In this manuscript, we offer an extra dimension to the evidence base for immediate decision-making, as well as anticipating future information from the immunological research community about the relationship between SARS-CoV-2 exposure and immunity.

## Materials and Methods

### Data Sources

We used publicly available epidemiological data to infer the underlying exposure to SARS-CoV-2 over time, as described below:

#### Regional daily death

The observed daily mortality data for each of 7 English regions (London, North West, North East (contains both North East and Yorkshire and the Humber regions), South East, South West, Midlands (West and East Midlands combined) and East of England) from January 1^st^ 2020 to November 11^th^ 2020 relates to daily deaths with COVID-19 on the death certificate by date of death. This information was extracted from the UK government’s official Covid-9 online dashboard^32^ on March 8, 2021.

#### Regional adjusted seroprevalence

Region specific SARS-CoV-2 antibody seroprevalence measurements, adjusted for the sensitivity and specificity of the antibody test, were retrieved from Public Health England’s National COVID-19 surveillance report^16^.

#### Regional case positivity ratios

Weekly positivity ratios of laboratory confirmed COVID-19 cases for each of 7 English regions were obtained from the week 40 and week 45 (2020) Public Health England’s National COVID-19 surveillance reports^16, 33^. The first report contains Pillar 1 testing information spanning weeks 5 (2020) to 39 (2020), and Pillar 2 positivity ratios from week 19 up to week 39 (2020). The second report presents both Pillar 1 and Pillar 2 testing data from week 27 to week 44 (2020). We took the average of both Pillar 1 and Pillar 2 test positivity ratios where both data were available.

#### Regional population

Region specific population structures were obtained from the UK Office for National Statistics 2018 population survey^34^.

### Mechanistic model

We developed a mechanistic mathematical model that relates reported COVID-19 daily deaths to seropositive status by assuming all COVID-19 deaths are reported and estimating an infection fatality ratio that is congruent with the observed seroprevalence data. For each region, *i* = 1, …, 7 corresponding to London, North West, North East, South East, South West, Midlands and East of England respectively, we denote the infection fatality ratio at time *t* by *α*_*i*_(*t*) and the number of daily deaths by *m*_*i*_(*t*). While we formulate the model in terms of a general, time-dependent, infection fatality ratio, we assume its default shape to be time invariant and later allow infection fatality ratio to vary with the stage of the epidemic.

Using the diagram in Figure 1 as reference, and given a number of observed deaths at time *t, m*_*i*_(*t*), we can expect a number of infections 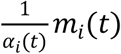 to have occurred *d*_*e*_ days before. Of these infected individuals, *m*_*i*_(*t*) will eventually die, whilst the remaining 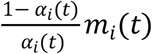 will seroconvert from sero-negative to sero-positive. This assumes that seroconversion happens, on average, with the same delay from the moment of infection as death.

Assuming that seropositive individuals convert to seronegative (serorevert) at a rate *β*, the rate of change of the number of seropositive individuals in region *i, X*_*i*_ *t*, is given by:

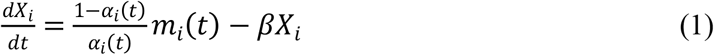

Solving Equation (1), subject to the initial condition *X*_*i*_ *t*_0_ = 0 where *t* is time since January 1st, 2020, gives:

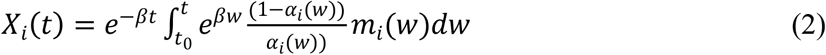

Discretizing Equation (2) with daily intervals (Δ*w* = 1) gives:

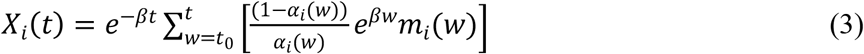

The model-predicted proportion of seropositive individuals in each population, *x*_*i*_(*t*), is calculated by dividing *X*_*i*_ (*t*) (Equation (3)) by the respective region population size at time *t*, 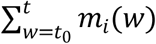, where *P*_*i*_ is the reported population in region *i* before the COVID-19 outbreak^34^:

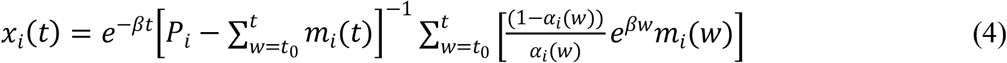

This is relatively straightforward when the serology data is already adjusted for test sensitivity and specificity as is the case. For unadjusted antibody test results, the proportion of the population that would test positive given the specificity (*k*_*sp*_) and sensitivity (*k*_*se*_) can be calculated as

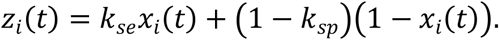

As mentioned earlier, the method that we present in this paper allows for the infection fatality ratio, *α*_*i*_(*t*), to be (a) constant or (b) vary over time with the stage of the epidemic:

a. For constant infection fatality ratio, we have:

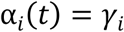
b. For time-varying infection fatality ratio, we first define the epidemic stage, *ES*(*t*), as the normalized cumulative positivity ratio:

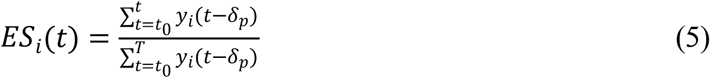

where *y*_*i*_(*t*) is the confirmed case positivity ratio at time *t* in the proportion of individuals testing positive for the virus, *δ*_*p*_ is the average time between testing positive and seroconversion (see Figure 1) and *T* is the total number of days from *t*_0_ until the last date of positivity data. In this work, we fixed *δ*_*p*_ = 7 days (see Figure 1 and main text). We assume that the infection fatality ratio is a linear function of the normalized cumulative positivity ratio as follows:

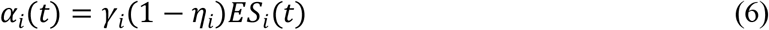

where *η*_*i*_ ∈ [0,1] and *γ*_*i*_ ∈ [0,1] are coefficients to be estimated. At the start of the epidemic when epidemic stage is 0 (see Equation (5)), then *α*_*i*_(*t*) = *γ*_*i*_, whereas when epidemic stage is 1, *α*_*i*_(*t*) = *γ*_*i*_ − *η*_*i*_ × *γ*_*i*_ ≤ *γ*_*i*_.

In Equation (5), *y*_*i*_(*t*) is taken from the regional weekly test positivity ratios (see Data section), converted to daily positivity ratios (taken to be the same over the week).

Once the model is parameterized, we can estimate the total proportion of the population that has been exposed, *E*_*i*_, with the following formula:

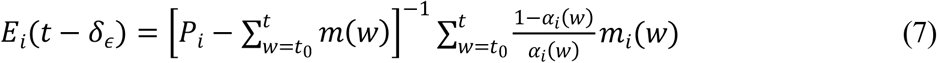

where *δ*_*ϵ*_ is fixed to 21 days (Figure 1).

### Observation model for statistical estimation of model parameters

We developed a hierarchical Bayesian model to estimate the model parameters *θ*, and present the posterior predictive distribution of the seroprevalence (Equation (4)) and exposure (Equation (7)) over time. Results are presented as the median of the posterior with the associated 95% credible intervals (CrI). We assumed a negative binomial distribution^35^ for the observed number of seropositive individuals in region *i* over time, 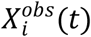:

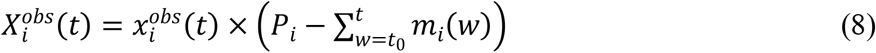

where 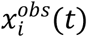 is the observed seroprevalence in region *i* over time. Then the observational model is specified for region *i* with observations at times 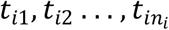:

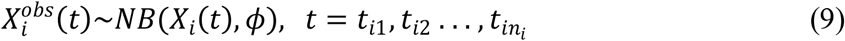

where *NB (X*_*i*_ *t, ϕ*) is a negative binomial distribution, with mean *X*_*i*_ *t* – given by equation (3)– and *ϕ* is an overdispersion parameter. We set *ϕ* to 100 to capture additional uncertainty in data points that would not be captured with a Poisson or binomial distribution. We assume uninformative beta priors for each of the parameters, according to the assumption made for how the infection fatality ratio is allowed to vary over time:

a. For constant infection fatality ratio, we have 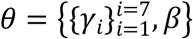 and take priors:

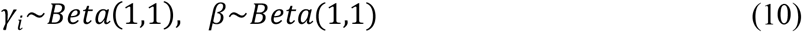
b. For time-varying infection fatality ratio, we have 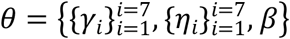 and take priors:

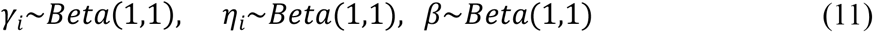

We use Bayesian inference (Hamiltonian Monte Carlo algorithm) in RStan^36^ to fit the model to seroprevalence data by running four chains of 20,000 iterations each (burn-in of 10,000). We use 2.5% and 97.5% percentiles from the resulting posterior distributions for 95% CrI for the parameters. The Gelman-Rubin diagnostics 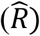 given in Table S1 and Table S

2 show values of 1, indicating that there is no evidence of non-convergence for either model formulation. Furthermore, the effective sample sizes (*n*_*eff*_) in Table S1 and Table S2 are all more than 10,000, meaning that there are many samples in the posterior that can be considered independent draws.

## Data Availability

All data, code, and materials used in the analyses can be accessed at:https://github.com/SiyuChenOxf/COVID19SeroModel/tree/master

## Acknowledgments

We would like to thank Adam Bodley for scientific writing assistance (according to Good Publication Practice guidelines) and editorial support. JF acknowledges funding from the Australian Research Council (DP200100747). LJW is funded by the Li Ka Shing Foundation, and the University of Oxford’s COVID-19 Research Response Fund (BRD00230). RA is funded by the Bill and Melinda Gates Foundation (OPP1193472).

## Author contributions

LJW and RA conceptualized the analysis; SC curated and formatted the data for analysis; SC and JAF developed the statistical model and performed the Bayesian inferences; LJW and RA wrote the first draft. SC, JAF, LJW and RA reviewed the results and edited the manuscript.

## Data and materials availability

All data, code, and materials used in the analyses can be accessed at: https://github.com/SiyuChenOxf/COVID19SeroModel/tree/master. All parameter estimates and figures presented can be reproduced using the codes provided. This work is licensed under a Creative Commons Attribution 4.0 International (CC BY 4.0) license, which permits unrestricted use, distribution, and reproduction in any medium, provided the original work is properly cited.

---

**Figure 2 — figure supplement 1.**
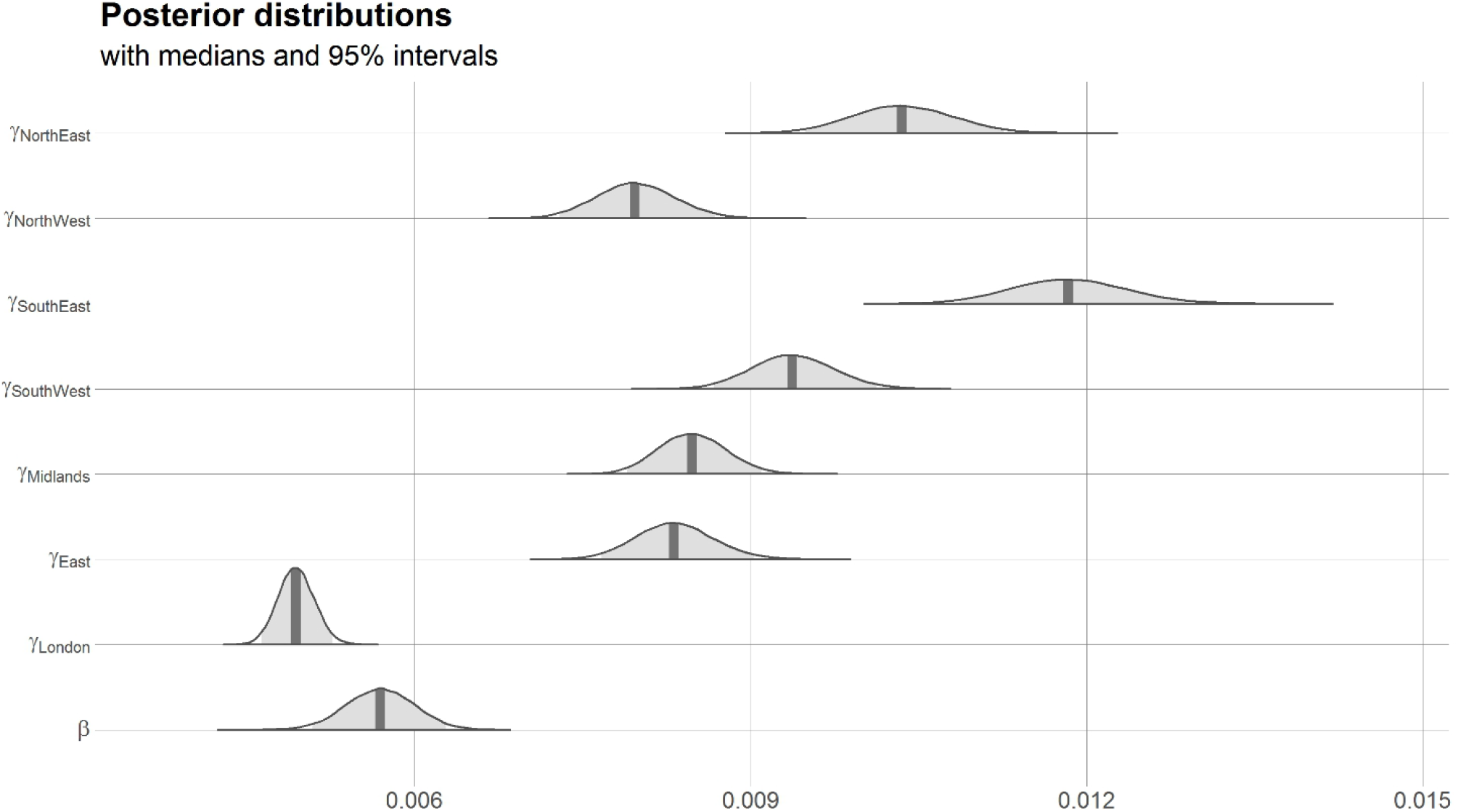
Marginal posterior distributions for parameters in the constant infection fatality ratio model. Vertical lines show median of distribution and grey shaded region shows 95% CrI.

**Figure 3— figure supplement 1.**
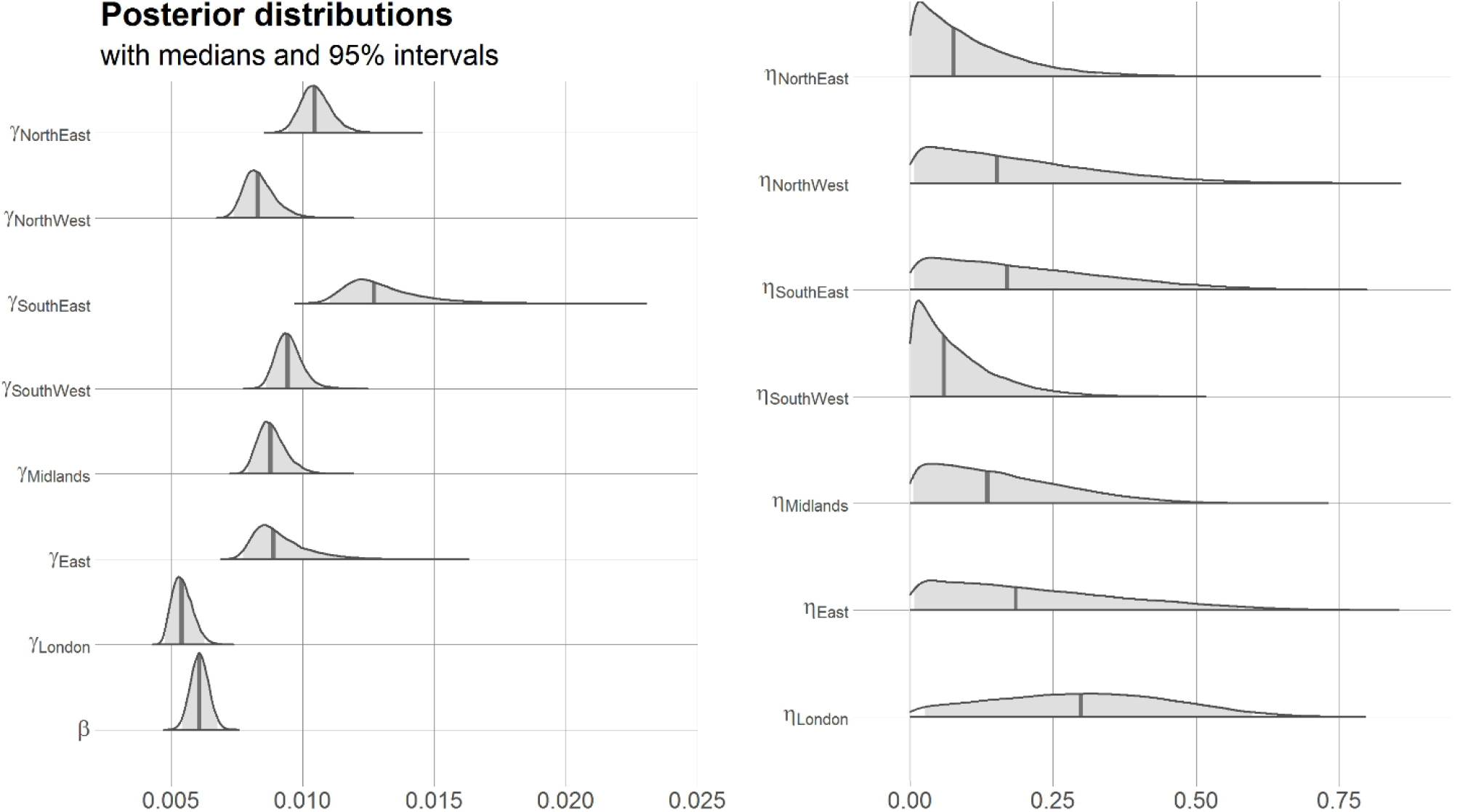
Marginal posterior distributions for parameters in the time-varying infection fatality ratio model. Vertical lines show median of distribution and grey shaded region shows 95% CrI.

**Figure 3– figure supplement 2.**
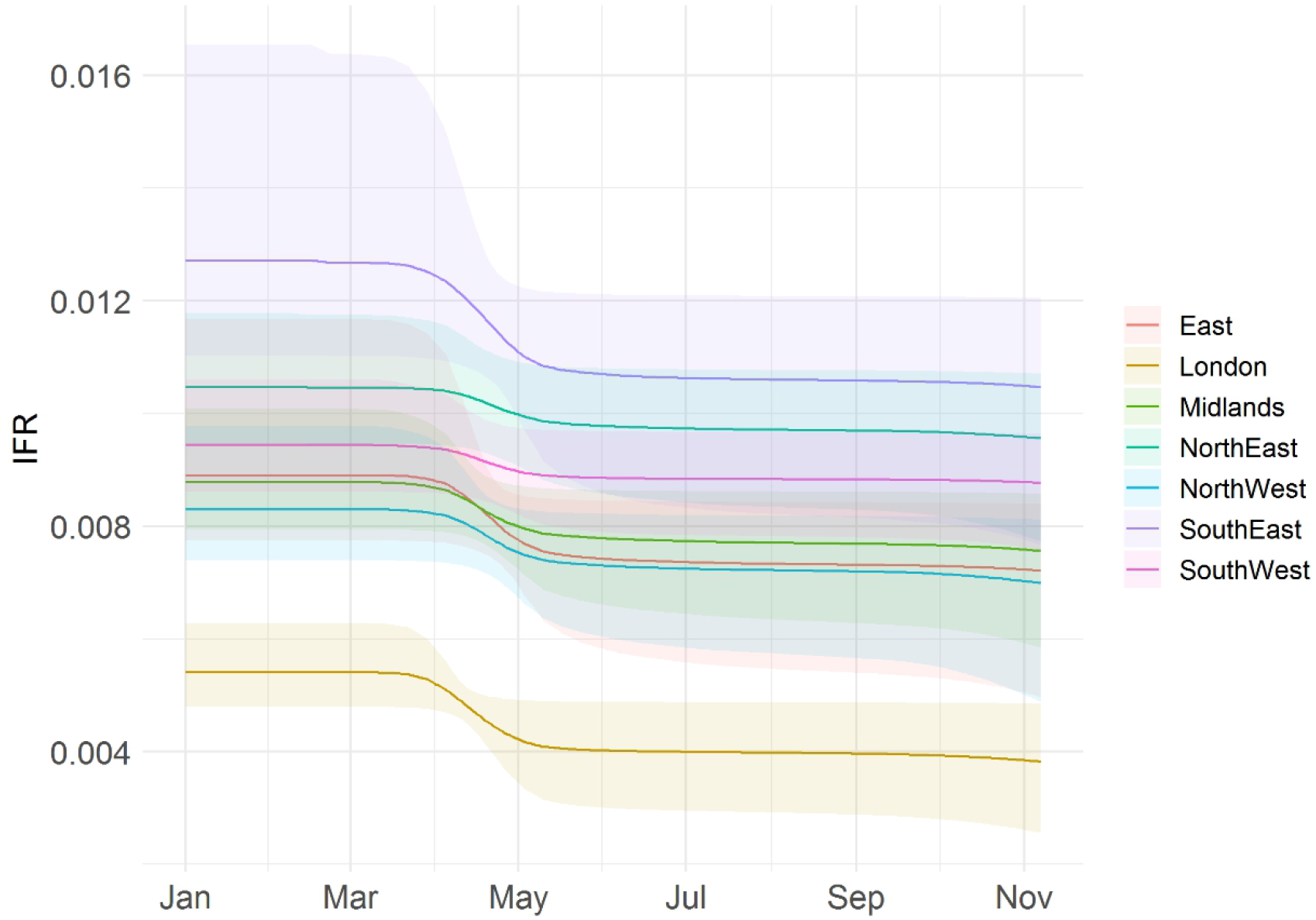
Posterior predictive distribution of time-varying infection fatality ratio. Solid line shows median and shaded region 95% CrI.

**Figure 3—figure supplement 3.**
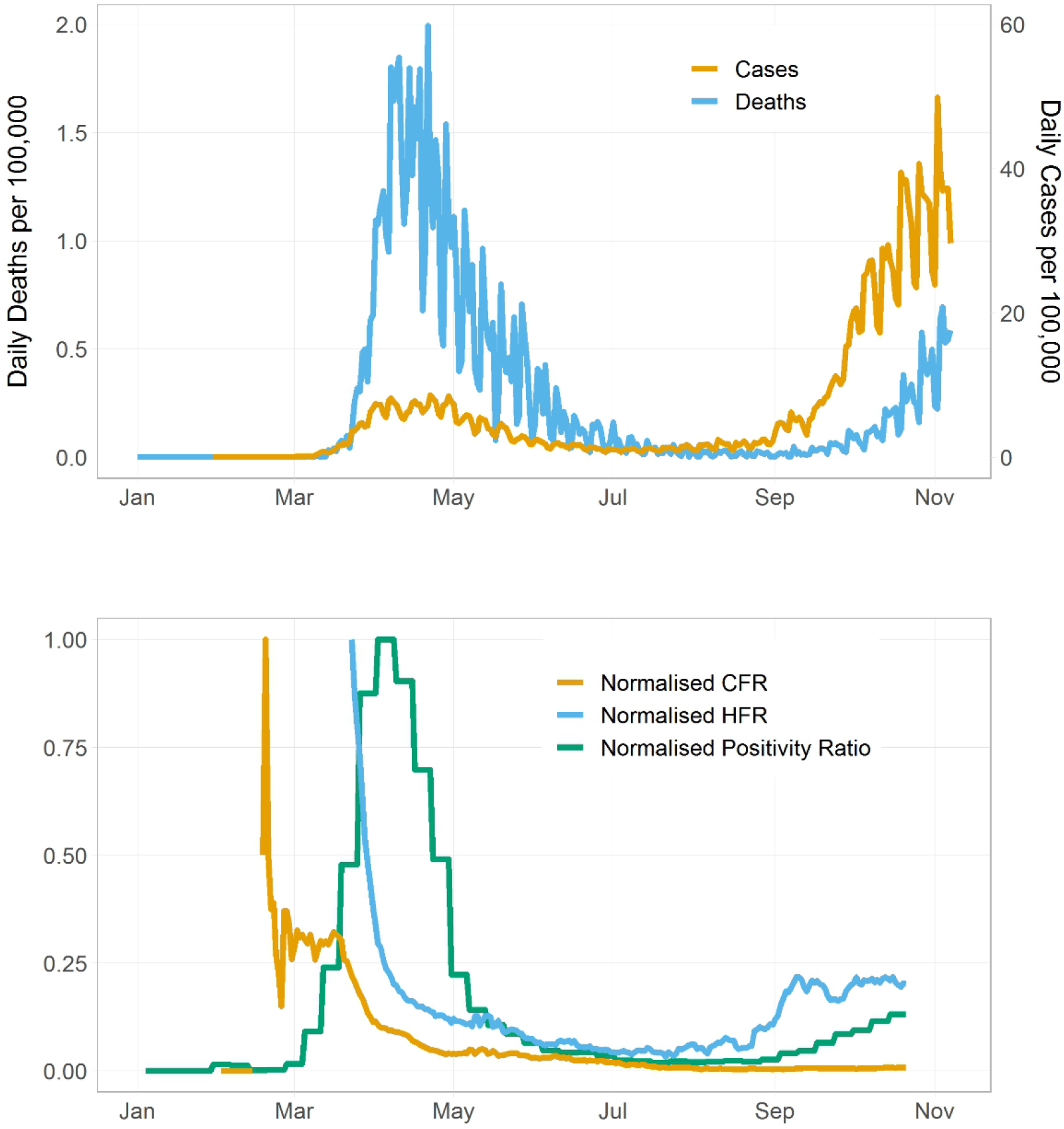
Relevant epidemiological metrics in England over the course of the pandemic. Top subplot shows COVID-19 cases and deaths (yellow and blue lines, respectively) per 100,000 population in England from Feb 5 2020 to Nov 7, 2020. Bottom subplot shows the normalized case fatality rate, hospital fatality rate and virus positivity (yellow, blue, and green lines, respectively). We assumed fixed time lags of *δ*_*b*_ = 14 days between PCR testing and death and *δ*_*h*_ = 12 days between PCR testing and hospitalization.

**Table S1.**
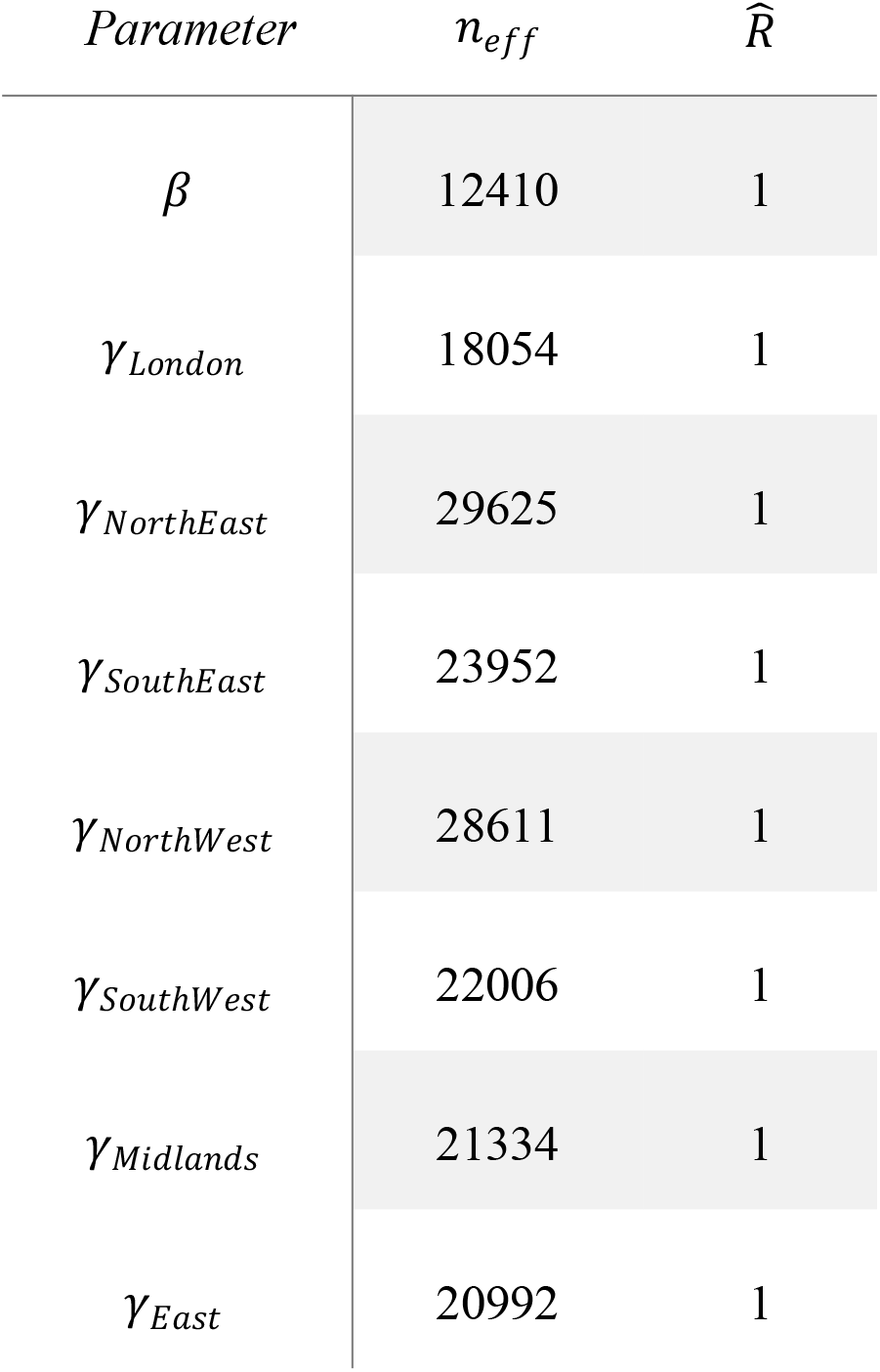
The effective sample size (*n*_*eff*_) and the Gelman-Rubin diagnostic 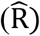 for the 8 model parameters in the default model (constant infection fatality ratio).

| *Parameter* | *n<sub>eff</sub>* | $\hat{R}$ |
| --- | --- | --- |
| $\beta$ | 12410 | 1 |
| $\gamma_{London}$ | 18054 | 1 |
| $\gamma_{NorthEast}$ | 29625 | 1 |
| $\gamma_{SouthEast}$ | 23952 | 1 |
| $\gamma_{NorthWest}$ | 28611 | 1 |
| $\gamma_{SouthWest}$ | 22006 | 1 |
| $\gamma_{Midlands}$ | 21334 | 1 |
| $\gamma_{East}$ | 20992 | 1 |

**Table S2.** The effective sample size (*n*_*eff*_) and the Gelman-Rubin diagnostic 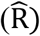 for the 15 model parameters in the time-varying infection fatality ratio model.

| *Parameter* | $n_{eff}$ | $\hat{R}$ |
| --- | --- | --- |
| $\beta$ | 35229 | 1 |
| $\gamma_{London}$ | 32184 | 1 |
| $\eta_{London}$ | 26680 | 1 |
| $\gamma_{NorthEast}$ | 38159 | 1 |
| $\eta_{NorthEast}$ | 33166 | 1 |
| $\gamma_{SouthEast}$ | 24036 | 1 |
| $\eta_{SouthEast}$ | 24430 | 1 |
| $\gamma_{NorthWest}$ | 32887 | 1 |
| $\eta_{NorthWest}$ | 29965 | 1 |
| $\gamma_{SouthWest}$ | 24430 | 1 |
| $\eta_{SouthWest}$ | 31047 | 1 |
| $\gamma_{Midlands}$ | 31943 | 1 |
| $\eta_{Midlands}$ | 27558 | 1 |
| $\gamma_{East}$ | 24703 | 1 |
| $\eta_{East}$ | 25506 | 1 |

**Table S3.** Marginal median parameter estimates and 95% CrI for the time-varying infection fatality ratio model.

| *Parameter* | *Median (95%CrI)* |
| --- | --- |
| $\beta$ | 0.0061 (0.0054 - 0.0068) |
| $\gamma_{London}$ | 0.0054 (0.0048- 0.0063) |
| $\eta_{London}$ | 0.30 (0.26 - 0.60) |
| $\gamma_{NorthEast}$ | 0.011 (0.0095 - 0.012) |
| $\eta_{NorthEast}$ | 0.077 (0.0029 - 0.33) |
| $\gamma_{NorthWest}$ | 0.0083 (0.0074 - 0.0098) |
| $\eta_{NorthWest}$ | 0.15 (0.0069 - 0.51) |
| $\gamma_{SouthWest}$ | 0.0094 (0.0086 - 0.011) |
| $\eta_{SouthWest}$ | 0.060 (0.0021- 0.26) |
| $\gamma_{SouthEast}$ | 0.0013 (0.011 - 0.017) |
| $\eta_{SouthEast}$ | 0.17 (0.0070 - 0.53) |
| $\gamma_{Midlands}$ | 0.0088 (0.0079- 0.010) |
| $\eta_{Midlands}$ | 0.14 (0.0060 - 0.42) |
| $\gamma_{East}$ | 0.0089 (0.0077 - 0.012) |
| $\eta_{East}$ | 0.18 (0.0080 - 0.57) |

